# Sepsis Transcriptomic Trajectory and Phenotypic Correlation Using Time Series Gene Expression from Childhood Meningococcal Infection

**DOI:** 10.1101/2022.11.10.22282189

**Authors:** Asrar Rashid, M. Toufiq, Praveen Khilnani, Zainab A. Malik, Z. Hussain, Hoda Alkhazaimi, Javed Sharief, Raziya Kadwa, Berit S. Brusletto, Amrita Sarpal, Damien Chaussabel, Rayaz Malik, Nasir Quraishi, Govind Benakatti, Syed A. Zaki, Rashid Nadeem, Guftar Shaikh, Ahmed Al-Dubai, Amir Hussain

## Abstract

Sepsis remains a leading cause of in-hospital morbidity and mortality. In the acute phase sepsis is described as a dysregulated process. The objective was to track temporal transcriptomic changes in infants with meningococcal disease (MSS). Therefore, two datasets underwent secondary analysis using temporal transcriptomic data. Methods applied to analysis included Principal Component Analysis (PCA), Transcript Time Course Analysis (TTCA) and Gene Set Expression Analysis (GSEA). Gene expression clustering algorithm for both datasets suggested three phases from the time of admission, and PCA plots indicated a gene-expression trajectory for both datasets. The data set from Kwan et al., showed that 410 genes differentiated survivors from the non-survivor, which included various cytokine, TNF, and apoptosis-associated pathways (normalized expression scores = −1.60, p = 0.02, and q = 0.15). Additionally, GSEA demonstrated gene sets significantly associated with disease severity. The genes for the cytokines CLC, HFE, HLA-F, NLRP3, and TNFRSF1B were significant. The Gene-sets elicited by GSEA in the Kwan and Emonts dataset had a high degree of molecular signature crossover (89.2% overlap), although a comparison of gene-expression trajectories across both datasets was difficult due to differing sample timing. Transcriptomic analysis demonstrated temporal trajectories in MSS-related gene expression. Larger studies using transcriptome expression are required could enhance the understanding of sepsis pathogenesis, improving prognostication and enable the development of precision therapies.

## Introduction

Pediatric and neonatal sepsis is an underestimated major global health problem. In 2017 sepsis in children constituted half the global sepsis burden, with 20 million episodes of pediatric sepsis and 2.9 million deaths in children younger than five years {Rudd, 2020 #3205}{WHO, 2017 #3396}. Children with sepsis have a mortality rate approaching 50% {Fleischmann-Struzek, 2018 #3397} and survivors have significant morbidity. Among children with severe sepsis that are referred to the Pediatric Intensive Care Unit (PICU), the majority of deaths occur within the first 24hrs from multi-organ failure or refractory shock {Cvetkovic, 2015 #3400}.

Early identification and timely intervention are key to improving clinical outcomes in children with suspected sepsis. Evidence-based guidelines have been developed to improve early recognition of sepsis using clinical and laboratory data {Weiss, 2020 #3267}{Casserly, 2015 #3407} {Goldstein, 2005 #1803}. Although adherence to sepsis guidelines and initiatives has improved, it has failed to translate into an improvement in mortality {Paul, 2018 #3409}{Larsen, 2021 #3410}. Early endotype prediction, such as using a biomarker risk model to predict children developing sepsis-associated myocardial dysfunction, could be useful for the application of precision strategies{Lautz, 2022 #4019}. Also, the clinical utility of measuring sepsis metabolites and biomarkers showed an improvement in the translation of cellular pathophysiology to clinical outcomes {Wang, 2020 #3411}{Kim, 2020 #3412}.

However, greater clarity is required to align the clinical features of the sepsis syndrome to altered immune function and physiological response, enabling more precise tailoring of treatment modalities. An enhanced understanding of sepsis dynamics is key to improving the prognostic capacity of biomarkers. Recently, a microarray-based approach utilizing messenger RNA (mRNA) gene expression has been used in sepsis prognostication and to develop precision therapies {Stanski, 2020 #2526}.

We hypothesized that microarray analysis of mRNA gene expression could have utility in better understanding the pathogenesis of Neisseria meningitides septic shock (MSS). We have analyzed differential mRNA gene expression in children with severe sepsis to establish a transcriptomic trajectory in patients with MSS to identify key time points for intervention to improve clinical outcomes.

## METHOD

### Public transcriptome dataset selection

Public, online repositories were searched to find clinical studies involving temporal transcriptomic datasets involving Neisseria Meningococcal infection. The search term ‘Meningococcal Shock’ was parsed through the NCBI Gene Expression Omnibus (GEO) database (n= 64 datasets isolated) and Array Express EMBL-EBI (n=5 datasets isolated). Then the term ‘meningococcal sepsis’ was used in both Array Express (n=7 datasets isolated) and the NCBI GEO database (n=64 isolated). The term ‘meningococcal transcriptome’ was entered into Pubmed (n=8 datasets). Two publicly available datasets were finalized. Dataset descriptive information are shown with the two patient groups compared (Table 1). The Kwan dataset {Kwan, 2013 #49} provides the most frequent blood sampling in the first 48 hours of meningococcal sepsis, followed by the Emonts temporal data-set, which covers a longer period (72 hours) {Emonts Marieke, 2008 #2615}. In the Kwan dataset patient, 1 to 5 are denoted by P1 to P5, and in the Emonts dataset patient, 1 to 6 are represented by P1 to P6. The time points used in the study are given an arbitrary value according to research design, starting from time zero (T=0).

**TABLE 1:** Demographic Comparison for Kwan versus Emonts.

| Patient Identifier | 1 | 2 | 3 | 4 | 5 | 1 | 2 | 3 | 4 | 5 | 6 | *p-Value Comparison |
| --- | --- | --- | --- | --- | --- | --- | --- | --- | --- | --- | --- | --- |
| Mortality (at 28 days) | Died | Alive | Alive | Alive | Alive | Alive | Alive | Alive | Alive | Alive | Alive |  |
| Neisseria Meningococcal Serotype | Negative culture (presumes meningococcal sepsis) | GpB | GpB | GpB | GpB | GpB | GpB | GpB | PCR negative | GpB | GpB |  |
| Title | 1N | 2N | 3N | 4N | 5N | 1R | 2R | 3R | 4R | 5R | 6R |  |
| DIC | Y | Y | Y | Y | Y | Y | Y | N | Y | Y | N |  |
| Mechanical Ventilation | Y | Y | Y | Y | Y | Y | Y | Y | Y | Y | Y |  |
| Gender | Female | Female | Female | Male | Male | Male | Male | Male | Male | Male | Male |  |
| Protein C | Y | N | N | N | N | N | N | N | N | N | N |  |
| Hospital | Kwan | Kwan | Kwan | Kwan | Kwan | Emonts | Emonts | Emonts | Emonts | Emonts | Emonts |  |
| Number of samples taken | 5 | 5 | 5 | 5 | 5 | 4 | 4 | 4 | 4 | 4 | 4 |  |
| Age (years) | 1.08 | 0.83 | 1.83 | 2 | 0.75 | 5.06 | 1.37 | 1.79 | 2.09 | 1.51 | 8.04 | 0.13 |
| Duration of PICU admission (DAYS) | 9 | 4 | 3 | 6 | 3 | 4 | 5 | 1 | 2 | 60 | 4 | 0.46 |
| Median PRISM Score | 15 | 7 | 15 | 13 | 4 | 26 | 23 | 21 | 25 | 28 | 14 | 0.0035 |
| Leucocyte count (*10<sup>9</sup>/L) | 6.8 | 18.4 | 18.8 | 3.7 | 15.1 | 16.1 | 5.1 | 35.5 | 18.1 | 8.3 | 9.9 | 0.6 |
| Platelet count (*10<sup>9</sup>/L) | 129 | 126 | 123 | 88 | 82 | 111 | 26 | 184 | 91 | 116 | 124 | 0.97 |
| CRP (mmol/L) | 52 | 81 | 60 | 138 | 96 | 51 | 78 | 176 | 258 | 105 | 31 | 0.44 |
| Lactate (mmol/L) | 6.75 | 0.99 | 1.71 | 6.48 | 0.6 | 9 | 5.7 | 1.7 | 1.8 | 9.5 | 2.2 | 0.43 |
| Base excess (mmol/L) | 1.2 | 2.4 | -8 | -15.1 | -6.9 | -9 | -11 | -7 | -6 | -12 | -9 | 0.32 |
| Urea (mmol/L) | 11.7 | 4.9 | 8 | 6 | 3.3 | 7 | 7.4 | 4.4 | 11.4 | 7.6 | 6 | 0.77 |
| Potassium (mmol/L) | 4.5 | 4.5 | 4 | 3.4 | 3 | 3 | 3.7 | 3.2 | 3.4 | 3.2 | 3.4 | 0.14 |
| Bilirubin (micromol/L) | 12 | 7 | 5 | 3 | 4 | 6 | 9 | 3 | 3 | 12 | 8 | 0.78 |

\* A p-value comparison is shown comparing the Kwan group against the Emonts group, indicating that the two groups have similar demographic characteristics apart from the PRISM score.

### Analytical Strategy

The datasets were first inspected for data processing methods indicated by the author(s), and appropriate normalization and log2 transformation was applied wherever needed using R-script for further downstream analysis.

### Statistical and Gene Ontology analyses

Qlucore Omics Explorer (QOE) version 3.1 software (Qlucore AB, Lund, Sweden) was used to analyze the differential expression of the genes (DEGs). Box plots were used to illustrate differences in gene expression in QOE. GSEA uses a computational method to determine whether an apriori-defined set of genes show statistically significant differences between two biological states {Subramanian, 2005 #2568}. For GSEA, the mean expression value of multiple probe IDs that matched an official gene symbol was calculated. This value was considered to represent the expression intensity of the corresponding gene symbol. Hierarchical clustering was based on Euclidean distance and average linkage clustering.

Further, all gene expression data was centered to a mean equal to zero and scaled to a variance similar to one. A p-value of <0.05 was used to confirm statistical significance; a false discovery rate (FDR) or q value of < 0.25 was chosen for cut-off criteria. For GSEA, Gene ranks were downloaded from the broad institute database Molecular Signatures Database (MSigDB). In a ‘cross over’ GSEA method, we applied GSEA to the Kwan dataset to generate gene lists downloaded and checked for their presence in the Emonts dataset. ANOVA and t-testing was undertaken according to the design of the study sets. Further, GSEA-associated gene sets associated with cytokine production were used to analyze cell-signaling patterns in survivors versus the non-survivor.

### Transcript Time Course Analysis

Transcript Time Course Analysis (TTCA) R software allowed temporal data exploration {Albrecht, 2017 #2534}. In the Kwan dataset, the last sample (48-hour collection) was simulated as a constant, or control, using the last sample for each dataset. Emonts had age-gender matched controls. The KEGG database was used to obtain curated genes for Vascular Endothelial Growth Factor (VEGF) associated signaling, apoptosis, complement, and coagulation. The Hugo database was used to curate the Heat Shock protein genes, including HSBAP1, Zinc finger, and Metallothionein genes. Simple Over-representation analyses (via hypergeometric-distribution-based testing) was performed using the TTCA results. This provided an analysis of significant genes related to ‘Consensus,’ ‘Early Response,’ ‘Middle Response,’ ‘Late Response,’ ‘Complete Response,’ ‘Dynamic,’ and ‘MaxDist.’ Similarly, pathfindR software {Ulgen, 2019 #3312} was used to analyze the genes from the TTCA. KEGG pathway and Gene Ontology Biological Process (GO-BP) enrichment analysis was undertaken using enrichR {Kuleshov, 2016 #3313}.

## Result

The Kwan dataset allowed a comparison of gene expression according to survival. The statistical filtering led to the isolation of 410 genes differentiating the survivors from the non-surviving patient. ICAM3 expression differed significantly according to survival (p=1.895e-05). CRP did not show a similar differentiation. When GSEA was applied to differentiate according to disease severity, a gene signature for GOBP was identified for altered cytokine production. Of this 88 gene signature set, five genes (CLC, HFE, HLA-F, NLRP3, TNFRSF1B) were present in the Kwan dataset.

### Emonts Data Analysis across different cell sub-types

The Emonts dataset includes data from peripheral mononuclear blood cells (PMBCs), lymphocytes, and monocytes and included control samples (Figure 1C).

**Figure 1.**
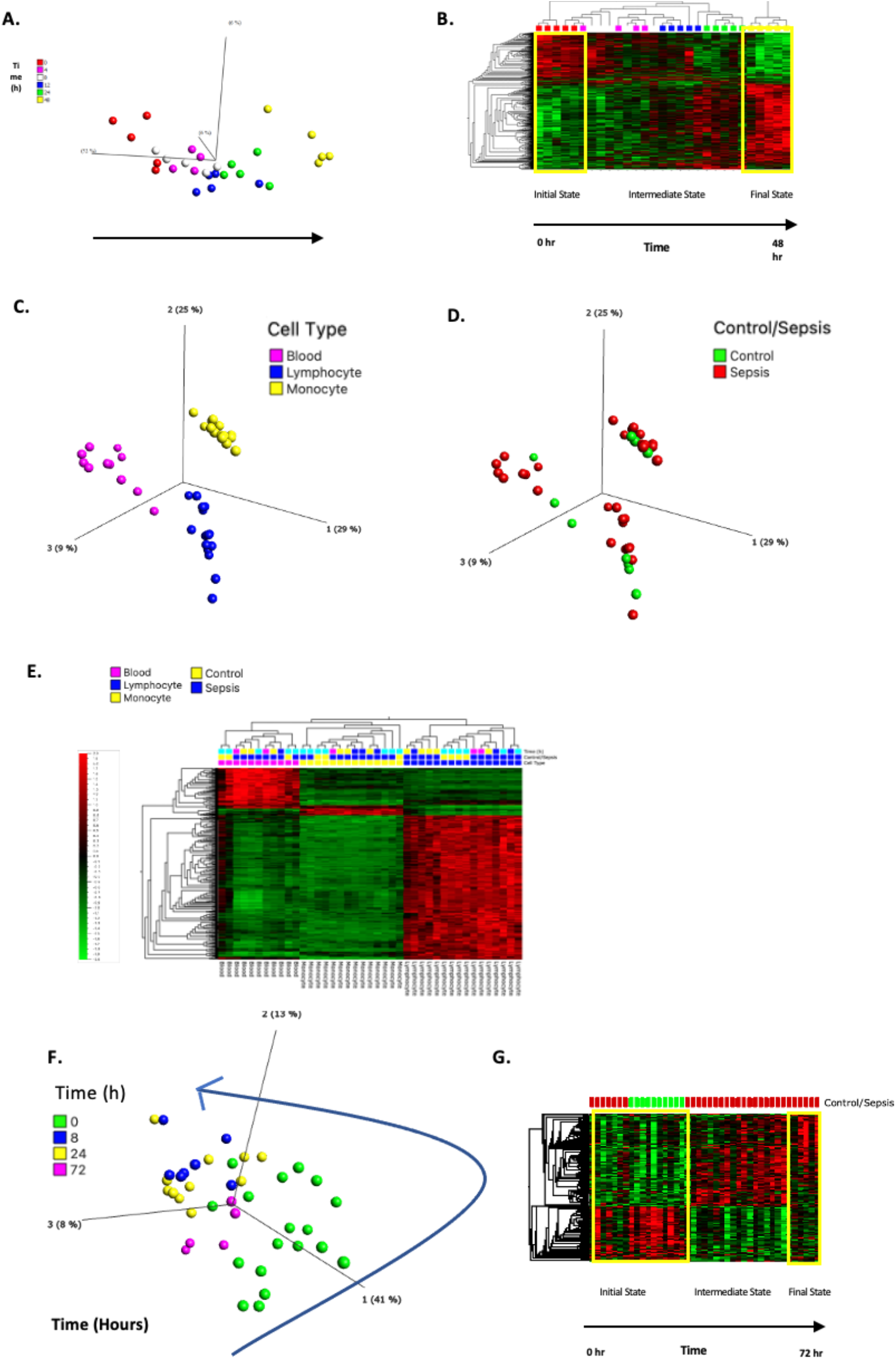
Gene Expression Trajectory from 2 data series of infants with Meningococcal Sepsis. Figure **1A**: Kwan data-set Multi-group analysis (p≤0.01, 728 genes) showing the expression levels of 728 genes define differences between time points. The PCA plot shows the time as being the single most important component in differences between samples (principal component, 52%). The Kwan dataset elicited 728 genes associated with time as the independent variable. Figure **1B:** Heat map with red indicating up-regulation of gene expression, green indicating low expression. At time 0, the majority of these genes exhibit low expression, by 48h, the majority are increased (see yellow boxes). The heap map shows that gene expression transitions from an initial state to a final state. The vertical and horizontal hierarchical clustering heat-map indicated that time points 0 and 48 hours were the most different in terms of gene expression and demonstrated the greatest transition from one pattern to another. Emonts Data PCA is shown according to different cell types (Blood, Lymphocyte and Monocytes), ANOVA undertaken (p=0.01 and q=0.012, giving 4543 genes of significance (Figure **1C**: ANOVA according to cell type p=0.01 and q=0.012). Controls are included in this dataset (Figure **1D**). PCA is from a Multi-group ANOVA analysis with respect to Time (eliminating by ANOVA the ‘Cell type”), (p=0.002, q=0.14, axis 41%, 13%, and 8%), 776 genes were differentially up-regulated across time points (Figure **1E**) with the associated heatmap (Figure **1F)** which again shows the transition from an initial state to a final state.

### Time Series Analysis

ANOVA PCA analysis of gene expression datasets from infants with MSS in the Kwan and Emonts studies identified a trajectorial pattern (Figures 1A and 1D). This was supported by heat maps (Figures 1B and 1E) demonstrating a starting state,

### Kwan data set according to survival

PCA was used to compare gene expression in relation to survival (Figure 2A), and the fatal case (P1) showed differing gene expression compared to the survivors (P2-P5). Gene sets related to various cytokines, TNF, and apoptosis were used for GSEA (Figures 2 E & F).

**Figure 2.**
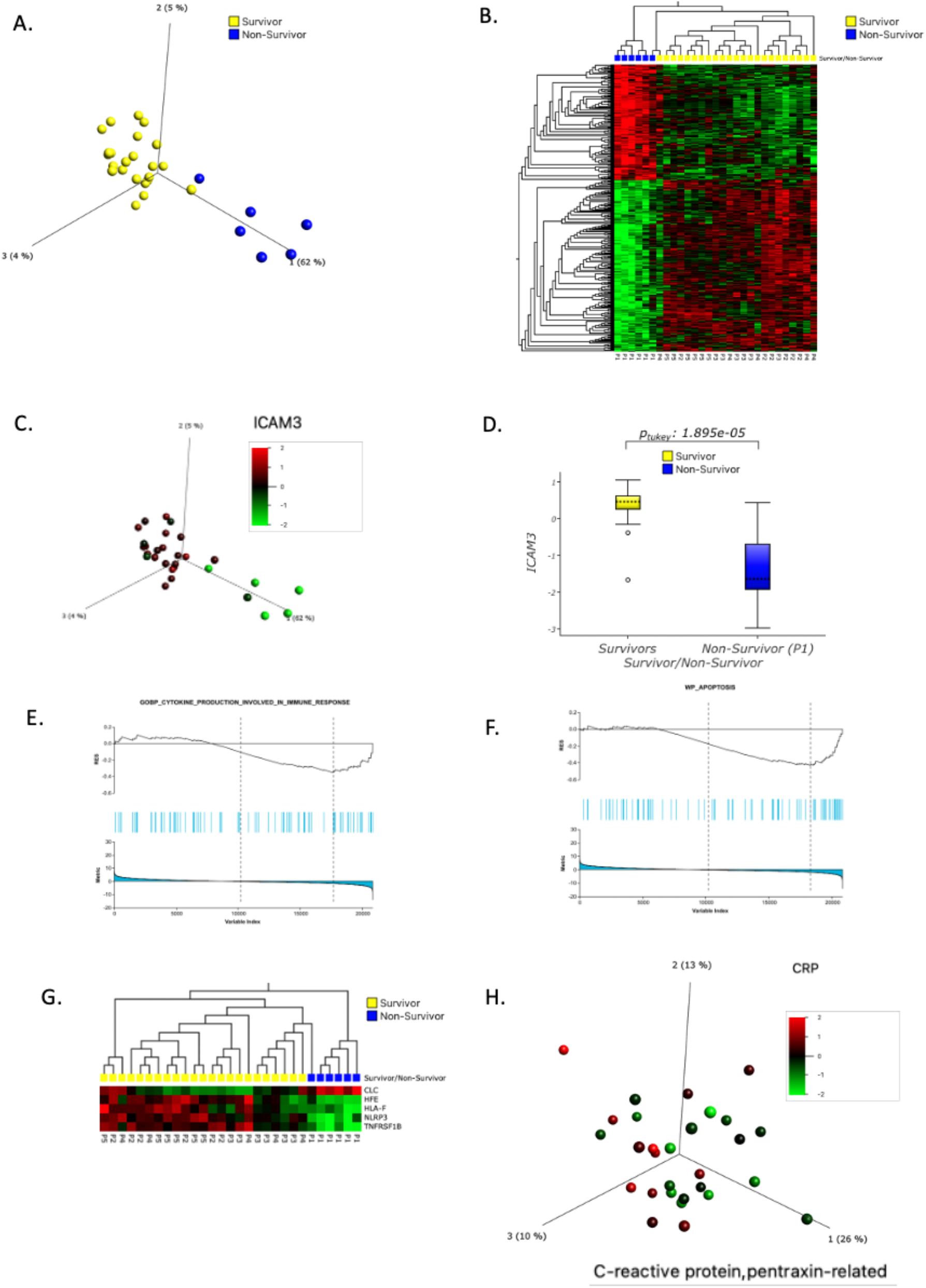
Kwan, non-survivor versus survivors, differential gene expression, heat plot, and GSEA. Figure **2A** **Kwan survival analysis**. 2 group (t-test) by ‘Patient ID,’ P1 (non-survivor) versus the surviving patients (P2-P5) elicited 410 genes that differentiated the two group PCA (p=1.0e-04, q=0.008), in the non-collapsed gene symbol mode (Figure **2A)**. The differential gene expression between the two groups is shown in the form of the heat-map, the vertical axis representing the 410 genes (Figure **2B)**. ICAM3 was included in the list of 410 genes and shown here with a heat map, showing up-regulation in the surviving patients (Figure **2C**). GSEA was undertaken, t-Test ‘Patient ID,’ P1 (non-survivor) versus the surviving patients (P2-P5). Figure **2E:** GSEA was undertaken using cytokine.gmt, GSEA significant P1 versus the other patients for GO_BP_CYTOKINE_PRODUCTION_INVOLVED_IN_IMMUNE_RESPONSE and WP_APOPTOSIS Figure **2F**. The Gene-set GO_BP_CYTOKINE_PRODUCTION_INVOLVED_IN_IMMUNE_RESPONSE contained 83 genes and was checked in the original 410 gene list. Five genes (genes CLC, HFE, HLA-F, NLRP3, TNFRSF1B) overlapped both this molecular gene set and the original 410 genes (Figure **2G)**, differentiating the non-survivor (P1) against the surviving patients as one gene probe occurred twice in the gene list, the gene-symbols were collapsed according to averaging. Gene expression for C-reactive protein was not seen to differentiate from a survival point-of-view (Figure **2H**) (Pro-Calcitonin gene was not available on this microarray panel).

A. The cytokine gene set differentiating P1 from P2-P5 included the ‘ALTEMEIER_RESPONSE_TO_LPS_WITH_MECHANICAL_VENTILATION’ pathway (NES=-1.68, p=0.004 and q = 0.14) and ‘GO_BP_CYTOKINE_PRODUCTION_INVOLVED_IN_IMMUNE_RESPONSE’ pathway (NEW=-1.53, p=0.022, q= 0.24)

Negative enrichment here implies genes from the Kwan et al. dataset were downregulated against the MSigDB geneset.

B. Apoptosis.gmt for P1 versus P2-P5 gave a gene set related to WP_APOPTOSIS (NES=-1.61, p=0.02 and q=0.24). P2 versus P1,P3-P5 showed WU_APOPTOSIS_BY_CDKN1A_VIA_TP53 (NES=-1.89, p=0.02 and q= 0.06).

C. TNF related gene sets BIOCARTA_NFKB_PATHWAY (NES =-1.60,p=0.02,q=0.15) & ST_TUMOR_NECROSIS_FACTOR_PATHWAY (NES=-1.49, p=0.006 and q=0.08)

The gene set the GSEA (see above) for GO_REGULATION_OF_CYTOKINE_PRODUCTION_INVOLVED_IN_IMMUNE_RE SPONSE gene-list was overlapped into the Kwan Data (Figure 1E).

### Comparing Kwan and Emonts datasets using GSEA

GSEA was also used to compare the Emonts and Kwan datasets (Table 2). First, GSEA was applied to the Kwan dataset eliciting significant gene-lists using ANOVA, ‘Time (hours),’ using Hallmark genes (H.aal.v7.2 symbols GMT). Gene overlap was assessed according to gene symbol by performing a cross-over comparison of significant gene lists and showed a high percentage overlap between the two independent datasets. Herein gene lists were highly conserved across the two datasets. GSEA was also applied in the Kwan dataset to compare the deceased patient with survivors (Table 3).

**Table 2:** Comparison of the Emonts versus Kwan Dataset using GSEA. Gene set generated from collapsing (average) the Kwan dataset and then applying Hallmark Gene Set for GSEA (h.all.v7.2.symbols.gmt), according to time. This led to column a. The data was then ported back into Emonts dataset (including controls and patients with MSS) to see the number of matches b. The percentage that matched in relation to the size of the original dataset are shown for Kwan (columns c) and for Emonts (column d). Enrichment score (ES), Normalised Enrichment score (NES), p and q values pertain to the GSEA undertaken on the Kwan dataset.

| Name of Gene Set | Size | Matches to the Kwan data-set<sup>a</sup> | Percentage Match to original gene list<sup>c</sup> | ES | NES | p value | q value | Matches to the Emonts dataset<sup>b</sup> | Percentage Match to original gene list<sup>d</sup> |
| --- | --- | --- | --- | --- | --- | --- | --- | --- | --- |
| HALLMARK_MYC_TARGETS_V1 | 200 | 185 | 92.5% | 0.53 | 1.98 | 0.002 | 0.03 | 191 | 95.5% |
| HALLMARK_BILE_ACID_METABOLISM | 112 | 111 | 99.1% | 0.40 | 1.37 | 0.003 | 0.21 | 112 | 89.2% |
| HALLMARK_COAGULATION | 138 | 132 | 95.7% | 0.39 | 1.40 | 0.007 | 0.21 | 138 | 100% |
| HALLMARK_XENOBIOTIC_METABOLISM | 200 | 190 | 95.0% | 0.40 | 1.41 | 0.007 | 0.23 | 195 | 97.5% |
| HALLMARK_CHOLESTEROL_HOMEOSTASIS | 74 | 69 | 93.2% | 0.50 | 1.62 | 0.007 | 0.16 | 71 | 95.9% |
| HALLMARK_MTORC1_SIGNALING | 200 | 189 | 94.5% | 0.46 | 1.67 | 0.01 | 0.19 | 195 | 97.5% |
| HALLMARK_HYPOXIA | 200 | 193 | 96.5% | 0.39 | 1.40 | 0.015 | 0.20 | 194 | 97% |
| HALLMARK_REACTIVE_OXYGEN_SPECIES_PATHWAY | 49 | 44 | 89.8% | 0.50 | 1.57 | 0.03 | 0.18 | 47 | 95.9% |

**Table 3:** t-Test Kwan dataset with one patient compared against all others in the group using GSEA. From the Kwan datasets Patients (P1 to P5) are compared against the collective using three sets of gmt files (Cytokine, TNF and Apoptosis). To right of each gmt classifier, the net enrichment scores (NES) breakdown according to sign is shown. A negative NES implies genes are down-regulated in the patient mentioned against the collective, whereas a positive sign suggests up-regulation of gene expression. On GSEA when the analysis does not generate genes there is no NES score i.e. Not Applicable (NA).

| Patient Against Rest | Cytokine.gmt | Negative or Positive NES scores | TNF.gmt | Negative or Positive NES scores | Apoptosis | Negative or Positive NES scores |
| --- | --- | --- | --- | --- | --- | --- |
| P1 versus Rest | 45 | All negative | 132 | All negative | 26 | All negative |
| P2 versus Rest | 6 | 1 negative, 5 Positive | 5 | All positive | 11 | 10 negative |
| P3 versus Rest | 2 | All positive | 0 | NA | 0 | NA |
| P4 versus Rest | 0 | NA | 0 | NA | 0 | NA |
| P5 versus Rest | 86 | 1 Negative, 85 Positive | 74 | All positive | 33 | All positive |
| GSEA Label | CYTOKINE | CYTOKINE | TNF | TNF | APOPTOSIS | APOPTOSIS |

### TTCA PathfindR-results

Data from the Kwan study was processed by TTCA and showed a decrease in pro-inflammatory IL1A and C3 compared to controls (Figures 3 A&G). TTCA also showed a slight increase then decrease in NFKB1 gene-expression, with an increase in TNF compared to controls (Figures 3 C&E). In the Emonts study TNF, IL1A and NFKB1 were increased compared to controls. Further, the Emonts dataset showed an initial fall of C3 gene expression between 0 to 24 hours, followed by a gradual rise (Figure 3H). Similarly, for the Emonts study, there was a fall in NFKB1 gene expression over the initial 8 hours, followed by a sustained rise.

**Figure 3.**
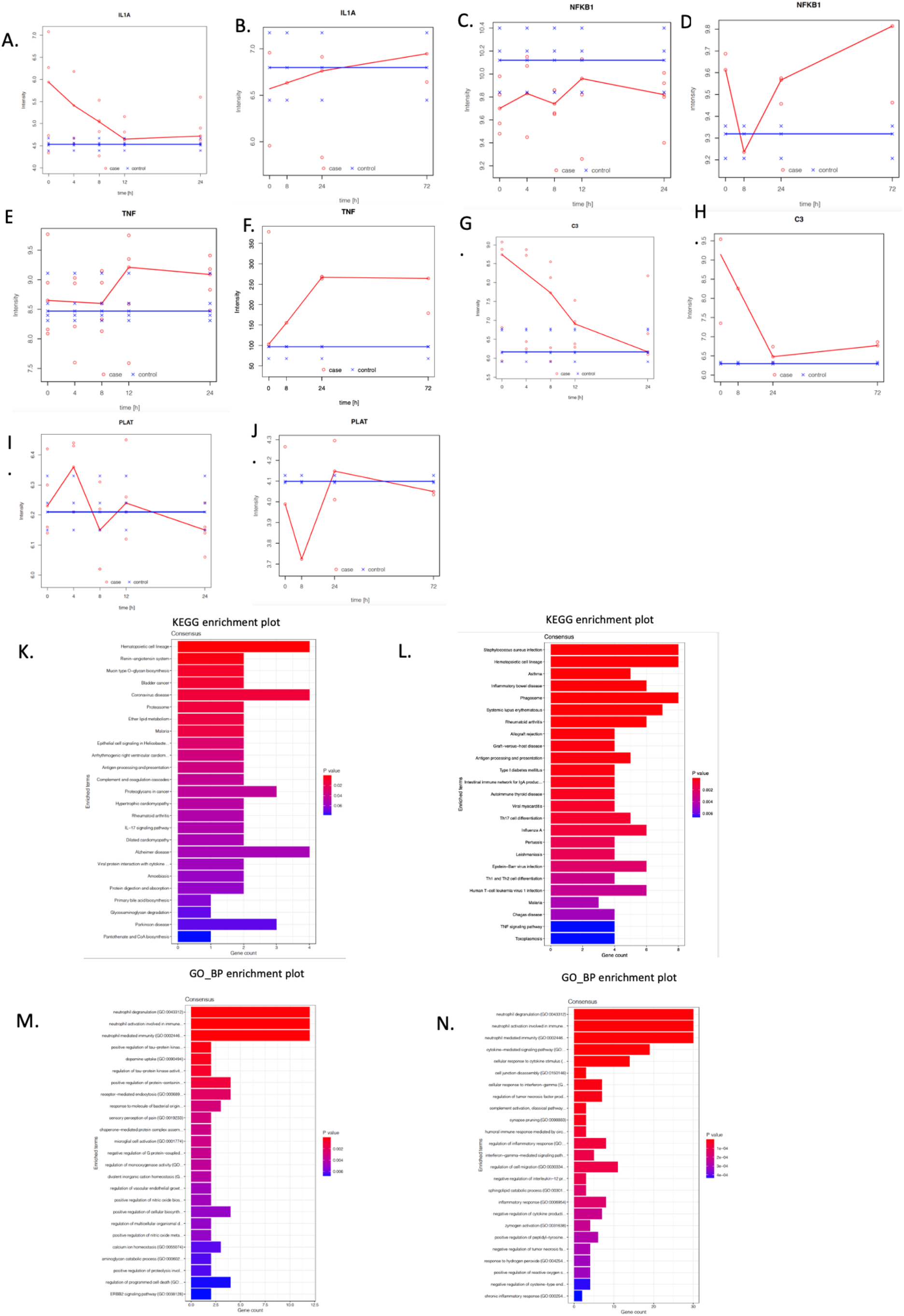
TTCA Analysis for the Kwan and Emonts Datasets. TTCA analysis for the Kwan dataset (Figure **3 A, C, E, G**, and **I**) and the Emonts dataset (Figure 3 **B, D, F**, and **H**). Using data from TTCA, data bar plots were generated for Kwan (Figure **3 K** and **M**) and Emonts (Figure **3 L** and **N**). Using TTCA, IL1A shows a fall for Kwan (Figure **3A**) but a rise in Emonts (Figure **3B**). NFKB1 a fall and climb over the 24 hours (Figure **3C**), whereas a rise is noted in Kwan (Figure **3D**). TNF a rise is noted in both datasets (Figure **3E** and **F**), C3 a fall in both datasets (Figure **3G** and **H**), and PLAT a rise and fall noted in both (Figure **3I** and **J**). KEGG enrichment shows the coronavirus pathway to be enriched for both datasets (**K** and **L**), antigen presentation, and hematopoietic cell lineage.

Comparison across the datasets was difficult due to the differing time periods of both studies. IL1A gene expression decreased from 0 to 8 hours and then showed a positive linear gradient from 12 to 24 hours in the Kwan dataset, whereas a consistent increase occurred from 0 to 72 hours in the Emonts study. There was a similar rise and fall of NFKB1 in both datasets. However, the intensity scales (y-axis are) differ. The shape of the TNF intensity and fall of C3 gene expression in one dataset by TTCA was similar to the other datasets. The Kwan and Emonts datasets have a similar shape for C3 gene expression from 0-24 hours, with a continuing increase in the Emonts study from 24 to 72 hours. A zig-zag change in intensity occurred for PLAT gene expression with a fall between 12 to 24 hours for Kwan and 24 to 72 hours for Emonts datasets.

Curated genes from TTCA analysis were enriched using PathfindR with enrichment charts eliciting significant pathways (Figure 4). Further, for both datasets, enrichment plots showed significance for the hematopoietic cell lineage (3 J&K), and GO_BP enrichment plots showed significance for the neutrophil degradation pathway (Figure 3 L&M). KEGG gene enrichment showed the presence of pathways related to Coronavirus infection, Ribosome, and th17 cell differentiation in both the Kwan and Emonts datasets. Table 4 shows TTCA gene enrichment comparison of Kwan versus Emonts datasets.

**Table 4:**
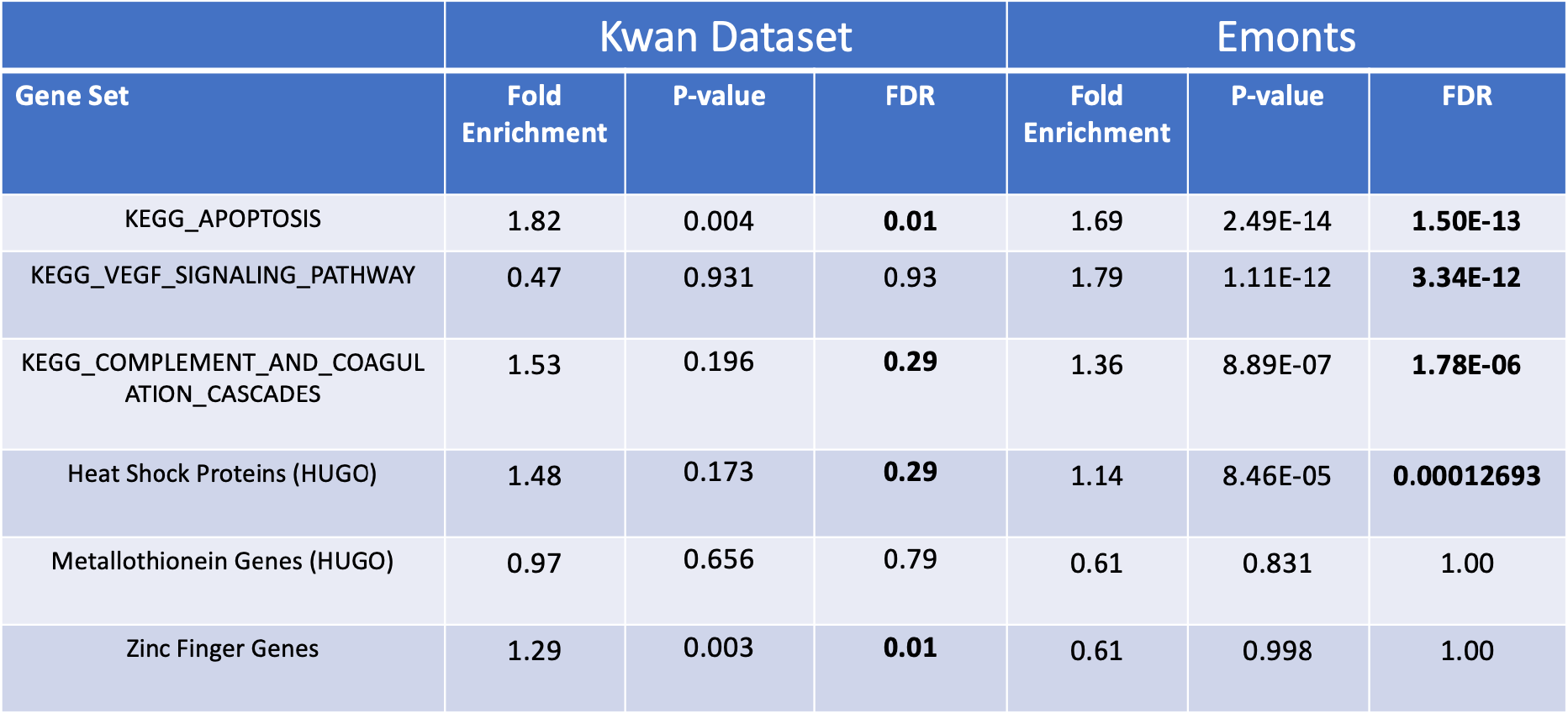
Curated genes from TTCA gene enriched according to the ‘Complete Response’ category. TTCA analysis and enrichment generated the ‘Complete Response’ category for both the Emonts and the Kwan Datasets. The pathway gene sets are compared for each dataset. Genes are enriched for apoptosis in both datasets according to the KEGG apoptosis pathway. For Kwan the Zinc Finger Genes are enriched, whereas not for Emonts. For Emonts VEGF, complement and coagulation cascades and Heat Shock Proteins are also noted.

| Gene Set | Kwan Dataset |  |  | Emonts |  |  |
| --- | --- | --- | --- | --- | --- | --- |
|  | Fold Enrichment | P-value | FDR | Fold Enrichment | P-value | FDR |
| KEGG_APOPTOSIS | 1.82 | 0.004 | **0.01** | 1.69 | 2.49E-14 | **1.50E-13** |
| KEGG_VEGF_SIGNALING_PATHWAY | 0.47 | 0.931 | 0.93 | 1.79 | 1.11E-12 | **3.34E-12** |
| KEGG_COMPLEMENT_AND_COAGULATION_CASCADES | 1.53 | 0.196 | **0.29** | 1.36 | 8.89E-07 | **1.78E-06** |
| Heat Shock Proteins (HUGO) | 1.48 | 0.173 | **0.29** | 1.14 | 8.46E-05 | **0.00012693** |
| Metallothionein Genes (HUGO) | 0.97 | 0.656 | 0.79 | 0.61 | 0.831 | 1.00 |
| Zinc Finger Genes | 1.29 | 0.003 | **0.01** | 0.61 | 0.998 | 1.00 |

**Figure 4:**
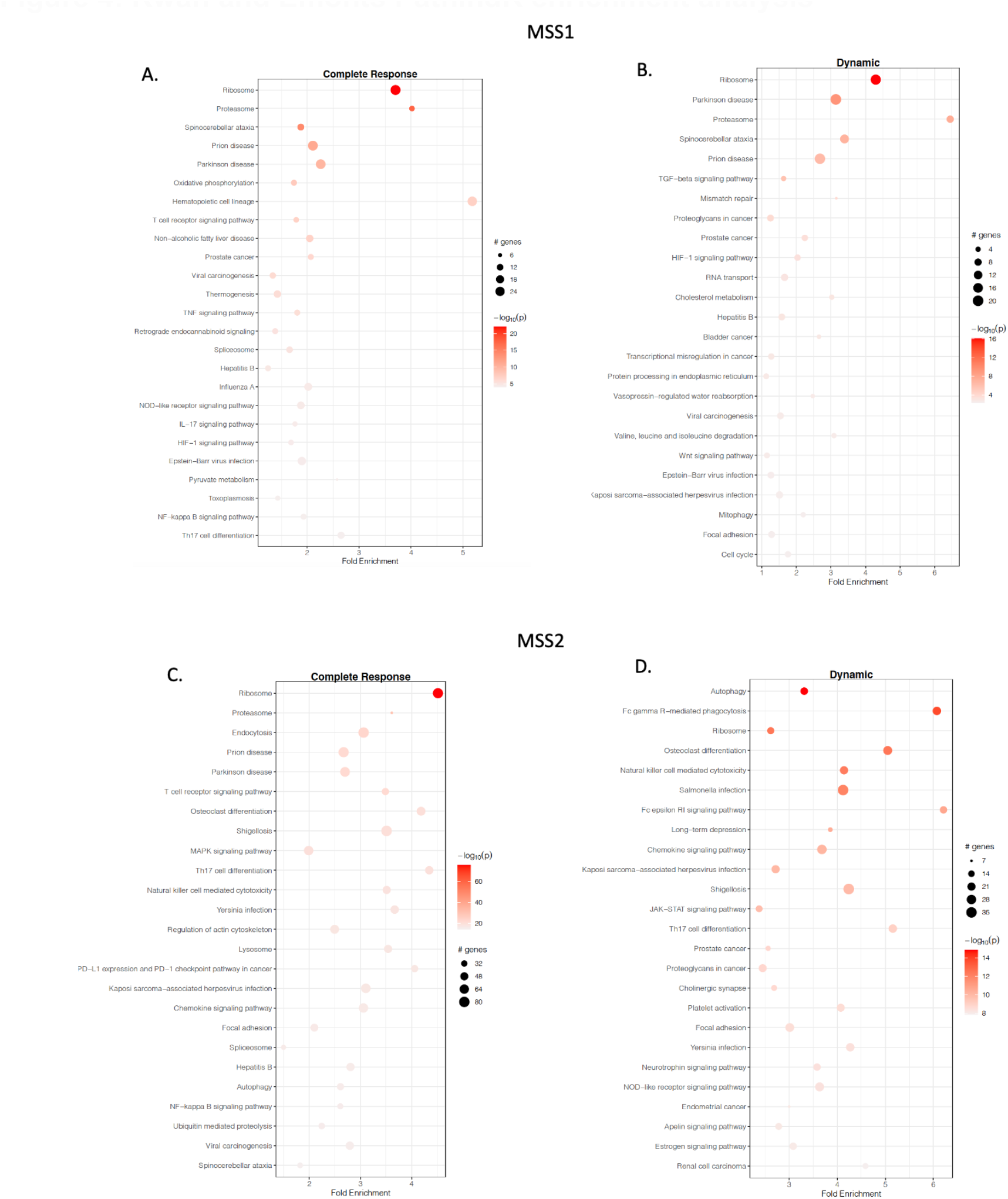
Kwan and Emonts PathindR enrichment analysis. From the TTCA data, PathindR enrichment charts were generated for both the Kwan (Figure 4**A** and **B**) and Emonts datasets (Figure 4**C** and **D**). For both datasets, Ribosome gene enrichment was significant for complete and dynamic responses. Also, T cell receptor signaling pathways were noted to be significant in the complete response for both datasets. Th17 cell differentiation was noted to be significant in Kwan (complete response) and Emonts (complete and dynamic response).

## Discussion

Severe sepsis has been regarded as a dysregulated immune process. Thereby the primary aim of this study was to enhance our understanding of the immune repertoire during severe sepsis by utilizing mRNA-based analysis. Two independent clinical temporal datasets in infants with acute invasive meningococcal disease were explored regarding gene expression. Data was filtered to differentiate genes and discern their relationship to time, allowing a dynamic representation of altered gene expression. PCA and heat map plot analysis depicted three phases in the trajectorial patterns. Also, PCA of the Kwan data separated gene expression according to survival. Therefore a gene-expression trajectory suggests a degree of temporal immune organization in severe sepsis, at least at the level of transcript signaling.

GSEA aids the interpretation of differences in molecular signatures from gene expression datasets. Thus using GSEA, Kwan et al. showed a temporal fall in oxidative phosphorylation gene expression {Raman, 2015 #42}. Further GSEA-based modular cluster analysis has shown value in identifying early predictive biomarkers of sepsis {Li, 2021 #3099}{Zeng, 2021 #3503}, relating changes to outcomes by Kaplan-Meier analysis {Groeneveld, 2019 #3501}. A novel application of GSEA in the current study was to identify differences between survivors and non-survivors with MSS. Furthermore, on labeling the Kwan data according to gene symbol, Hallmark gene sets showed molecular signatures of significance. Moreover we showed that these gene sets were also identifiable in the Emonts dataset. Suggesting a high degree of conservation for molecular signatures across MSS datasets. Such GSEA methodology may enhance the understanding of dynamic alterations in molecular signatures and could aid in prognostication.

The Kwan et al. dataset contained a list of genes potentially useful to differentiate survivors from non-survivors, including ICAM-3, NLRP3 and TNFRSF1B. Differential host gene expression analysis showed ICAM3 gene up-regulation after 6 hours. Of relevance, ICAM3 is expressed in a human meningeal-based model incubated with meningococci, indicating the importance of ICAM-3 in invasive meningococcal disease {Robinson, 2004 #3492}. ICAM-3 may have a role in early antigen-independent T-cell interactions with antigen-presenting cells, which may be necessary for pathogen surveillance {Montoya, 2002 #3493}. Its role in severe sepsis and up-regulation associated with mortality has not been previously documented. Although ICAM3 has been shown to bind to the dengue virus envelope protein and given the endothelialopathy in Dengue, the predilection for endothelial damage in MSS could be mediated via ICAM3 {Kerrigan, 2019 #3494}. Garnacho-Montero et al. showed elevated NLRP3 gene expression in critically ill patients, particularly those with sepsis, and this was sustained today 7 in sepsis survivors {Garnacho-Montero, 2020 #3496}. Our study indicates an initial rise in NLRP3 in the fatal case compared to the survivors in the first 48 hours. NLRP3 is an inflammasome that activates macrophages, epithelial cells, and endothelial cells to produce pro-inflammatory cytokines IL-18 and IL-1β, coined the ‘cytokine storm’ in response to the host-pathogen {Fricke-Galindo, 2021 #3499} and was first popularized with H5N1 influenza (Avian) virus infection {Tisoncik, 2012 #3498}. The NLRP3 inflammasome pathway also has significance for SARS-CoV2 disease and is responsible for the leukopenia, neutrophilia, and inflammation associated with COVID-19 {Fricke-Galindo, 2021 #3499}{Farahani, 2021 #3500}. TNFRSF1B gene expression was increased in the non-survivor. This gene is a part of the gene-expression profile in a murine model of sepsis-associated pulmonary dysfunction {Wang, 2018 #3082}. TNFRSF1B has also been expressed in vivo and in vitro following influenza virus infection with a pulmonary cytokine storm {Wang, 2018 #3082}. Also, both datasets confirmed proof of common mechanistic pathways with coronavirus through TTCA analysis. TTCA allowed a temporal illustration of gene expression and was combined with Pearson correlation and pathR enrichment analysis. While there were potential similarities in trajectories on TTCA, this was difficult to quantify due to the differing design of the two independent studies. Further, identifying KEGG gene enrichment pathways relating to coronavirus infection suggests an overlap in disease pathogenesis between MSS and Severe Acute Respiratory Syndrome (SARS).

This analysis has limitations such as the fact that the Kwan cohort lacked controls which would have been useful in mitigating confounding factors. A work around in TTCA analysis was to labele the last Kwan sample in the time series as the control, assuming that the child was then the most stable. In contrast, the Emonts dataset had age and gender-matched controls, which possibly resulted in the different significant gene sets generated by TTCA compared to the Kwan data. Also, the small sample numbers in both studies is a limitation as well as the differences in the duration of study between the two datasets. The Kwan dataset was derived over 48 hours compared to 72 hours in the Emonts study. However, we believe that in the scientific literature no other clinical sepsis dataset currently surpasses the Kwan or Emonts datasets in terms of the number of transcript samples. We suggest that future studies should incorporate sequential sampling exploiting the temporal connectivity of data. However the optimal sample timing to define the transition of immunological function in acute sepsis remains unresolved. Kamaleswaran et al. described an Artificial Intelligence (AI) physiomarker multi-time point strategy for outcome prediction in sepsis{Kamaleswaran, 2018 #2671}. Such an AI-supervised approach combined with the temporal gene expression sampling could result in novel perspectives with respect to transcriptomic pathophysiology in acute severe sepsis.

### Conclusion

Transcriptomic analysis outlined in this paper has demonstrated three-phase temporal trajectories in MSS-related gene expression, with specific patterns related to survival. Future studies are warranted to validate the findings in this paper. Moreover, understanding how mRNA changes associated with MSS overlap with the cytokine storm described in SARS-CoV-2, requires further research.

## Data Availability

All data produced are available online at

## Data Availability

The secondary analysis of datasets undertaken in this paper were from gene expression microarray data available from public repositories. Readers can access the data from the online ArrayExpress database (EMBL-EBI ArrayExpress accession number E-MEXP-3850; https://www.ebi.ac.uk/arrayexpress/experiments/E-MEXP-3850) and from the National Library of Medicine GEO Dataset Repository ID GSE11755 https://www.ncbi.nlm.nih.gov/geo/query/acc.cgi?acc=GSE11755. There are no restrictions to data-access from the aforementioned repositories

## Acknowledgments

Dr David Inwald, PICU Addenbrookes Hospital, Cambridge, UK & To Professor Hector Wong for his great contribution to sepsis genomics

## Research and Ethics Committee

The paper was waived from review by the hospital Central Scientific Committee and the Research and Ethics Committee given the secondary analysis of data from primary studies.

## Author Contributions

Conceived and designed the experiments: AR Performed the experiments: AR

Analyzed the data: AR

Contributed analysis, methods, and tools: AR wrote the first draft of the paper: AR

Revised critically for importance and intellectual content: MT, PK, ZAM, ZH, JS, RK, BSB, AS, DC, RM, NQ, GB, SAZ, RN, GS, AA,AH

